# Common variation in a long non-coding RNA gene modulates variation of circulating TGF-*β*2 levels in metastatic colorectal cancer patients (Alliance)

**DOI:** 10.1101/2023.12.04.23298815

**Authors:** Julia C.F. Quintanilha, Alexander B. Sibley, Yingmiao Liu, Donna Niedzwiecki, Susan Halabi, Layne Rogers, Bert O’Neil, Hedy Kindler, William Kelly, Alan Venook, Howard L. McLeod, Mark J. Ratain, Andrew B. Nixon, Federico Innocenti, Kouros Owzar

**Author notes:** Correspondence: Kouros Owzar, PhD. Duke University School of Medicine. These authors contributed equally to this work (Federico Innocenti, Kouros Owzar).

## Abstract

**Background:** Herein, we report results from a genome-wide study conducted to identify protein quantitative trait loci (pQTL) for circulating angiogenic and inflammatory protein markers in patients with metastatic colorectal cancer (mCRC).The study was conducted using genotype, protein marker, and baseline clinical and demographic data from CALGB/SWOG 80405 (Alliance), a randomized phase III study designed to assess outcomes of adding VEGF or EGFR inhibitors to systemic chemotherapy in mCRC patients. Germline DNA derived from blood was genotyped on whole-genome array platforms. The abundance of protein markers was quantified using a multiplex enzyme-linked immunosorbent assay from plasma derived from peripheral venous blood collected at baseline. A robust rank-based method was used to assess the statistical significance of each variant and protein pair against a strict genome-wide level. A given pQTL was tested for validation in two external datasets of prostate (CALGB 90401) and pancreatic cancer (CALGB 80303) patients. Bioinformatics analyses were conducted to further establish biological bases for these findings.

**Results:** The final analysis was carried out based on data from 540,021 common typed genetic variants and 23 protein markers from 869 genetically estimated European patients with mCRC. Correcting for multiple testing, the analysis discovered a novel *cis*-pQTL in *LINC02869*, a long non-coding RNA gene, for circulating TGF-*β*2 levels (rs11118119; AAF = 0.11; *P*-value < 1.4e-14). This finding was validated in a cohort of 538 prostate cancer patients from CALGB 90401 (AAF = 0.10, *P*-value < 3.3e-25). The analysis also validated a *cis*-pQTL we had previously reported for VEGF-A in advanced pancreatic cancer, and additionally identified *trans*-pQTLs for VEGF-R3, and *cis*-pQTLs for CD73.

**Conclusions:** This study has provided evidence of a novel *cis* germline genetic variant that regulates circulating TGF-*β*2 levels in plasma of patients with advanced mCRC and prostate cancer. Moreover, the validation of previously identified pQTLs for VEGF-A, CD73, and VEGF-R3, potentiates the validity of these associations.

## Background

The heritability of circulating protein abundance and evidence showing the influence of germline genetic variants in circulating protein levels have raised the interest in protein quantitative trait loci (pQTL) studies. pQTL studies have the objective of determining the impact of germline genetic variants on circulating protein levels. Circulating protein levels are involved in diverse biological processes, including disease development and response to medications. pQTL analyses can contribute notably to the discovery of new clinically relevant biomarkers and to better understanding the factors that regulate circulating proteins and the pathways involved in these biological processes (1, 2).

Colorectal cancer (CRC) is the third most common type of cancer and the second leading cancer-related death worldwide (3). Many studies have attempted to identify circulating proteins as biomarkers in CRC patients, including biomarkers for the early detection of CRC (4, 5), prognosis (6, 7), treatment response (7), regional tumor localization (6), and disease dissemination (6). Thus, the assessment of the impact of germline genetic variants on circulating protein levels through pQTL analyses in CRC patients can potentially lead to insights into the mechanisms involved in CRC development and treatment outcome.

Herein, we report results from a study of common genetic variation with respect to variation in circulating proteins with putative inflammatory or angiogenic function in patients with metastatic (m) CRC. Specifically, we sought to identify functional *cis-* and *trans*-pQTL variants using genome-wide germline genotyping data and circulating protein levels measured using a custom panel of putative cancer-related angiogenic and inflammatory markers. To this end, we used clinical, genotyping, and pre-treatment candidate protein marker data obtained from patients with mCRC randomized to the Cancer and Leukemia Group B (CALGB, now part of the Alliance for Clinical Trials in Oncology (Alliance)) and the Southwest Oncology Group, CALGB/SWOG 80405. This was a phase III study randomizing mCRC patients to receive cetuximab, an epidermal growth factor receptor (EGFR) inhibiting monoclonal antibody, or bevacizumab, a vascular endothelial growth factor (VEGF) inhibiting monoclonal antibody, or the combination of the two in addition to systemic chemotherapy (8).

After accounting for multiple testing, our analysis discovered a novel *cis*-pQTL in the intronic region of *LINC02869* (alias *C1orf143*) for circulating TGF-*β*2 at the two-sided genome-wide level of 0.05. The novel *cis*-pQTL for TGF-*β*2 was then tested for validation in independent external cohorts of castration-resistant prostate cancer (CALGB 90401) and advanced pancreatic cancer patients (CALGB 80303) (9, 10, 11, 12, 13).

This analysis also validated a *cis*-pQTL for VEGF-A that we had previously identified in a cohort of patients with locally advanced or metastatic pancreatic cancer (13). Finally, our study identified *trans*-pQTLs for VEGF-R3 and *cis*-pQTLs for CD73.

## Methods

### Clinical data

Patients registered to CALGB/SWOG 80405 were randomized to receive bevacizumab, cetuximab, or the combination of these two monoclonal antibodies, in addition to systemic chemotherapy. For the latter, the choice of a FOLFOX- or FOLFIRI-based regimen was at the discretion of the treating physician. The study was later amended by restricting participation to patients with wild-type KRAS tumors and by terminating the combination arm. Additional details on the design of the study, its amendments, and clinical baseline characteristics and outcomes for its patients have been reported in a primary clinical publication and its supplementary material (8). Baseline demographic and clinical data used in the present analyses were obtained from the database used to generate the analyses reported in that publication.

### Genotyping data

Germline DNA was extracted from peripheral blood. The genotyping was conducted in two separate batches using the Illumina Human OmniExpress (12v1) and the Illumina Human OmniExpressExome (8v1) platforms, respectively, by the Core of Genomic Medicine of the RIKEN institute in Yokohama, Japan. The genotyping design included the use of HapMap controls as well as inter- and intra-plate replicates. The analysis data set was constructed on the basis of the intersection of the variants across these two platforms identified by their respective dbSNP Reference SNP id (rsid). A number of quality control (QC) metrics, including genotype calling rate, AAF, Hardy-Weinberg *P*-values, were used to filter out variants. Additional technical details on the genotyping and QC processes have been previously reported (14).

### Circulating protein markers

Levels of 23 soluble proteins (angiopoietin-2, HGF, ICAM-1, IL-6, OPN, PDGF-AA, PDGF-BB, PlGF, SDF-1, TGF-β1, TGF-β2, TIMP-1, TSP-2, VCAM-1, VEGF-A, VEGF-D, VEGF-R1, VEGF-R2, VEGF-R3, BMP-9, CD73, HER-3, TGFβ-R3) were measured in plasma from peripheral venous blood collected at baseline using multiplex enzyme-linked immunosorbent assay (ELISA). The plasma was double-spun, aliquoted, and frozen in liquid nitrogen. Additional technical details on this panel, including CVs, lower limits of quantitation, and limits of detection, have been previously reported (13, 15, 16, 17). The analyses reported herein are based on measurements taken at baseline prior to any CALGB/SWOG 80405 protocol-directed treatment.

### Statistical considerations

To ensure robustness against outliers and influential data points and deviations from normality assumptions, the Jonckheere-Terpstra statistic (18, 19) was used for the discovery of pQTLs. Then variance approximation provided in expression 6.19 in Hollander *et al.* (20), implemented by the fastJT package (21), was used to derive a standardized statistic whose null sampling distribution was approximated using a standard normal distribution.

To properly account for multiple testing in the discovery process, a conservative two-sided genome-wide significance level of 0.05/*K*, where *K* denotes the number of SNP and protein marker pairs tested in the final analysis, was used. The potential confounding effects of baseline covariates, including age at time of registration (log base 10 transformed), self-reported gender, and global ancestry, was assessed using a robust linear regression rank-based approach implemented by the Rfit (22) package. The genotype effect was quantified on the additive scale as the number of copies of the alternate allele (additive genetic model), and global ancestry was inferred for pateints previously identified as genetic Europeans (14) using the first three principal components estimated using the SNPRelate R package (23). The Hodges-Lehmann-Sen estimator was used to estimate the location parameter for the distribution of the abundance of a protein conditional on the genotype. The per allele effect size was estimated as the ratio of the location parameter estimates. A 95% exact confidence interval was calculated for each location parameter. These estimates were meant to serve as descriptive measures, and accordingly, the corresponding confidence levels were not adjusted for multiple testing. For each protein marker, the distribution of the unadjusted *P*-values was examined using Manhattan and QQ plots.

All statistical analyses were conducted using the R statistical environment (24) and its extension packages, including those from the tidyverse (25) ecosystem, foreach (26), SeqArray(27), kableExtra (28), knitr (29) and rmarkdown (30). SNP and gene positions are reported per GRCh37.

### Replication analysis

For a given pQTL pair, the Jonckheere-Terpstra statistic (18, 19) with the genotype effect quantified on the additive scale as the number of copies of the alternate allele (additive genetic model) was used to estimate the pQTL association in two independent external datasets, CALGB 90401 (9) and CALGB 80303 (11, 13). CALGB 90401 included metastatic castration-resistant prostate cancer randomized to receive docetaxel in combination with prednisone on day 1 plus either placebo or bevacizumab every 21 days. CALGB 80303 included patients with advanced pancreatic cancer randomized to receive gemcitabine on days 1, 8, and 15 plus either placebo or bevacizumab on days 1 and 15. Additional details on the design of both studies, and clinical baseline characteristics and outcomes for its patients have been reported in primary clinical publications (9, 11).

### Bioinformatics considerations

For a given pQTL pair, the extent of the signal, quantified by unadjusted *P*-values, relative to the positions of variants and their linkage disequilibrium (LD) within regions of annotated genes, was assessed visually using Locus Zoom ((31); version 1.4) plots. The June 2010 release The 1000 Genomes Pilot 1 EUR panel (November 2014; hg19 coordinates using GENCODE gene annotation (32)) was used as the reference. Putative functional effects were investigated using RegulomeDB (33), USCS Genome Browser (34), Haploreg (35), and SNPNexus (36). AtSNP was used to quantify the impact of SNPs on transcription factor binding (37).

## Results

The final analysis was conducted on the basis of a data set comprised of 540,021 single nucleotide polymorphisms (SNPs), 23 baseline protein markers, and baseline demographic and clinical data from 869 genetically estimated European mCRC patients from CALGB/SWOG 80405 for whom protein marker data was available. The Consolidated Standards of Reporting Trials (CONSORT (38)) chart displayed in Innocenti, *et al.* (14) provides additional details on the sample and variant selection process leading to the final analysis data set. **Table 1** provides summaries of baseline demographic and clinical data for the analysis cohort of this study. The two-sided genome-wide level significance threshold was set to be 4.03e-09. At this level, 37 candidate pQTLs across four proteins, TGF-*β*2, VEGF-A, VEGF-R3, and CD73, were identified based on our pre-specified statistical decision rule. Overview and details for these candidates are illustrated in the Circos (39) plot in **Figure 1**, and summarized in **Supplementary Tables 1 and 2, Additional File 1**. The Manhattan and quantile-quantile (QQ)-plots for TGF-*β*2, VEGF-A, VEGF-R3, and CD73 are shown in **Supplementary Figures 1-4, Additional File 1,** respectively. Finally, for each of the 23 proteins, the top 100 pQTLs, ranked according to the corresponding unadjusted *P*-values are shown in **Additional File 2.**

**Figure 1.**
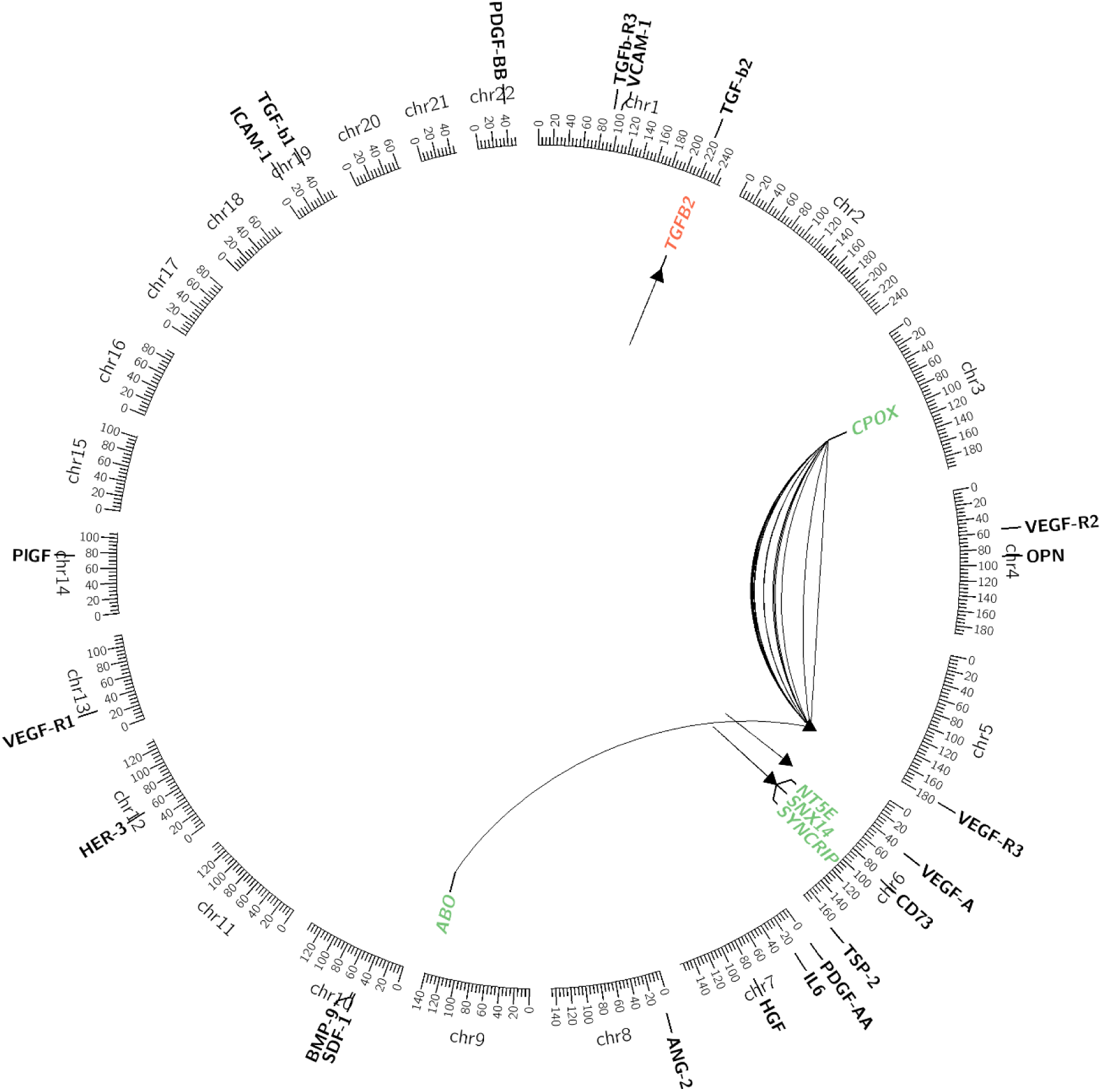
Chromosome-based Circos plot for pQTL that passed the genome-wide threshold. The colors indicate if a gene contains one of the top SNPs (green) or is a flanking gene (red). Links with less curvature indicate smaller *P*-values.

**Table 1.** Demographics and clinical characteristics of patients of genetically determined European ancestry included in the genome-wide pQTL analysis of CALGB/SWOG 80405.

|  | **Patients**<br>**n=869** |
| --- | --- |
| **Age (years) – median (range)** | 59.7 (21.8-85.3) |
| **Gender – n (%)** |  |
| Male | 506 (58.2%) |
| Female | 363 (41.8%) |
| **Treatment arm – n (%)** |  |
| Chemotherapy/Bevacizumab | 340 (39.1%) |
| Chemotherapy/Cetuximab | 312 (35.9%) |
| Chemotherapy/Bevacizumab/Cetuximab | 217 (25.0%) |
| **Chemotherapy – n (%)** |  |
| FOLFOX | 666 (76.6%) |
| FOLFIRI | 203 (23.4%) |
| **Prior adjuvant chemotherapy – n (%)** |  |
| No | 744 (85.6%) |
| Yes | 125 (14.4%) |
| **Prior pelvic radiation – n (%)** |  |
| No | 785 (90.3%) |
| Yes | 84 (9.7%) |
| **ECOG PS – n (%)** |  |
| 0 | 545 (62.7%) |
| 1 | 324 (37.3%) |
| **Tumor location – n (%)** |  |
| Left | 497 (57.2%) |
| Right | 230 (26.5%) |
| Transverse | 62 (7.1%) |
| Multiple | 4 (0.5%) |
| Unknown | 76 (8.7%) |

Our analysis identified a novel pQTL for TGF-*β*2: rs11118119 (chr1: 218693872; A>G; alternate allele frequency (AAF) = 0.11; *P*-value < 1.4e-14) is in the intronic region of *LINC02869* (alias *C1ord143*). This variant is located 75,911 bases downstream from *TGFB2*. The genotypic Hodges-Lehmann-Sen estimates are 95.8 (n = 692; 95% CI = 91.6, 100), 141 (n = 164; 95% CI = 128, 155) and 207 (n = 12; 95% CI = 140, 295) for genotypes AA, AG and GG respectively, while the median observed values were 87.2, 130.2, and 206.9, respectively. The estimated effect size in the rank-based linear model was 1.68 (CI = 1.51, 1.88). This variant is in moderate LD (R^2^ = 0.67 in the analysis data set) with rs725033 (chr1:218643940; G>A; AAF = 0.12; *P*-value < 1.6e-10) an intergenic variant 25,979 bases upstream from *TGFB2*. See **Table 2**, **Figure 2**, and **Supplementary Figures 5 and 6, Additional File 1** for box and locus zoom plots of rs11118119 and rs725033, respectively.

**Figure 2.**
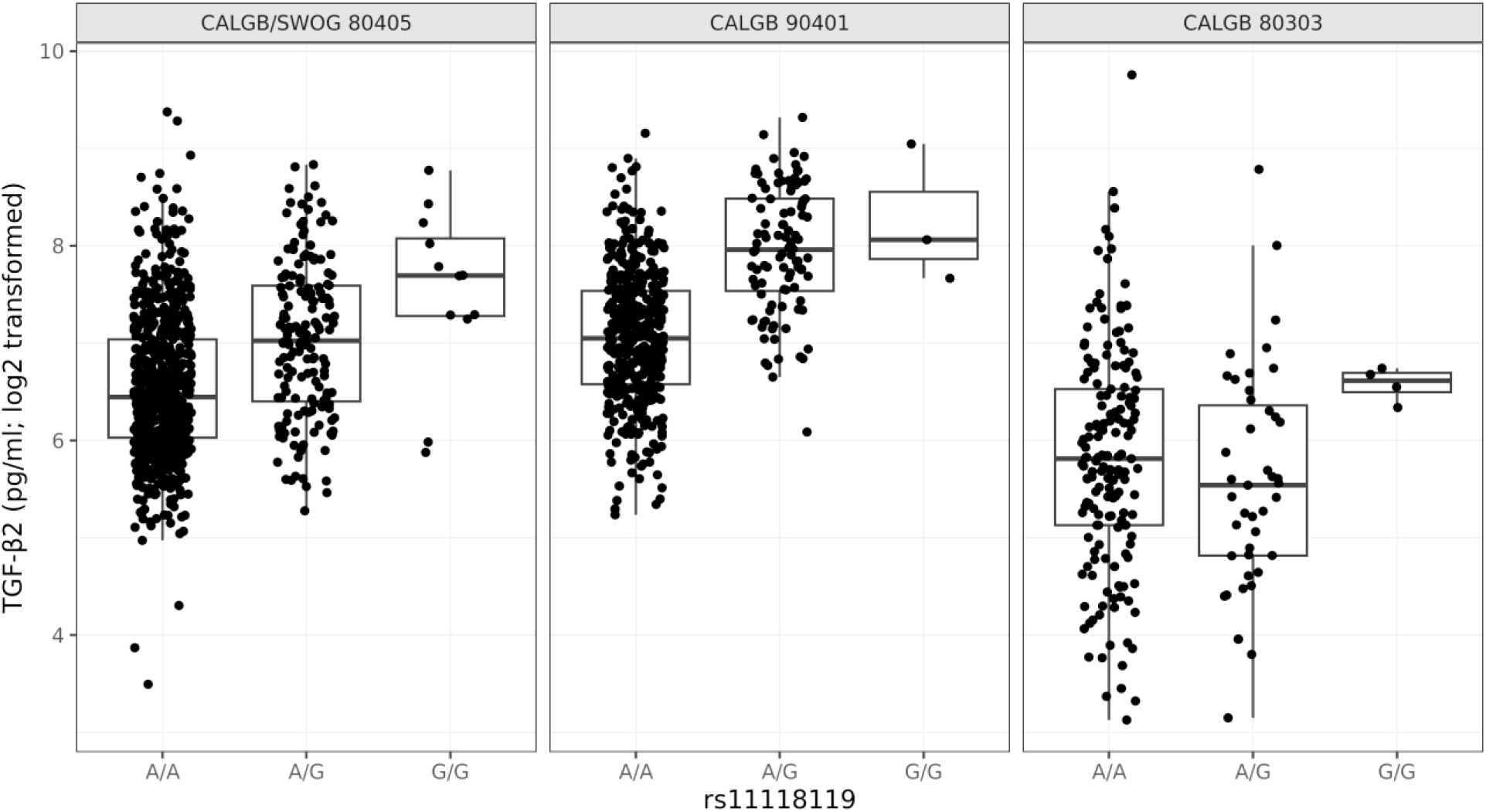
Associations between rs11118119 (A>G) and TGF-*β*2 levels in CALGB/SWOG 80405, CALGB 90401, and CALGB 80303.

**Table 2.**
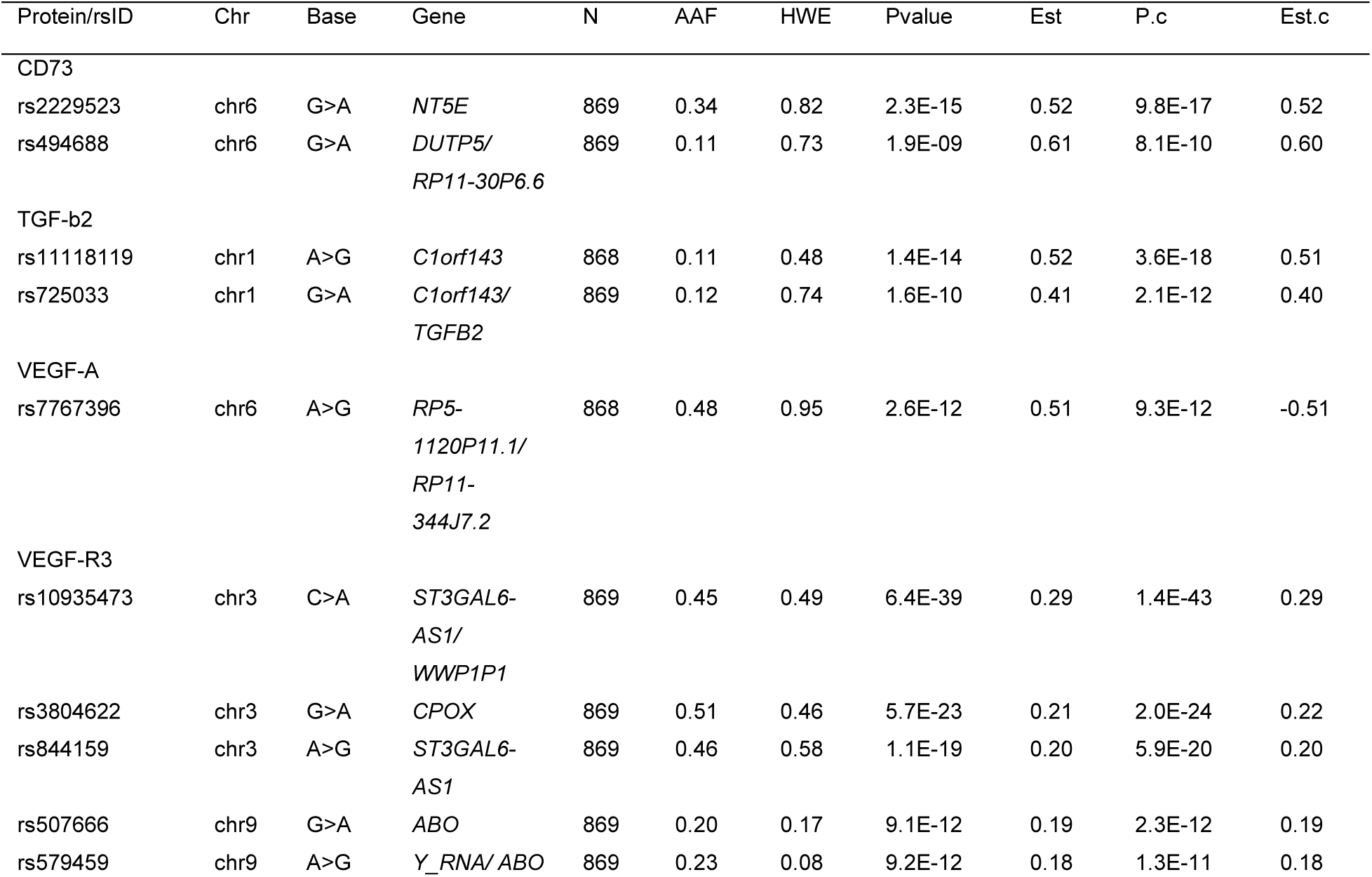
Results and annotation for pQTL that passed the genome-wide threshold. Protein/rsID: protein marker/RefSNP ID of variant; Chr: chromosome of the variant according to hg19; Base: Illumina TOP reference>alternate alleles; Gene: gene symbol for intragenic variants or nearest downstream/upstream gene symbols for intergenic variants; N: number of samples for whom the variant was called; AAF: relative allelic frequency for the alternate allele; HWE: Hardy-Weinberg *P*-value; Pvalue: *P*-value for pQTL analysis; Est: parameter estimate for the variant effect from the rank-based linear regression; P.c: *P*-value for the variant effect from the rank-based linear regression adjusting for covariates; Est.c: parameter estimate for the variant effect from the rank-based linear regression adjusting for covariates. Variants are sorted by rsID within Protein and Chr. Note that *C1orf143* is an alias for *LINC02869*, and that the genotyping data included rs3812138, but this ID has been merged with rs2229523.

Our analysis provided strong confirmatory evidence for a *cis*-pQTL we have previously reported for circulating VEGF-A in patients with locally advanced or metastatic pancreatic cancer: rs7767396 (chr6:43927050; A>G; AAF = 0.48; *P*-value < 2.6e-12), an intergenic variant 172,826 bases downstream from *VEGFA* and 41,272 bases upstream from *C6orf223*. See **Table 2**, and box and locus zoom plots in **Supplementary Figure 7, Additional File 1**.

Our analysis identified *trans*-pQTLs for VEGF-R3 on chromosomes 3 and 9. These include rs10935473 (chr3:98416900; C>A; AAF = 0.45; *P*-value < 6.4e-39), an intergenic variant 16,277 bases upstream from *ST3GAL6-AS1*, intronic variants in *CPOX* (e.g., rs3804622; chr3:98303182; G>A; AAF = 0.51; *P*-value < 5.7e-23), intronic variants in *ST3GAL6-AS1* (e.g., rs844159; chr3:98443648; A>G; AAF = 0.46; *P*-value < 1.1e-19) and an intronic variant in *ABO* blood group gene (rs507666; chr9:136149399; G>A; AAF = 0.20; *P*-value < 9.1e-12). See **Table 2**, and **Supplementary Figures 8, 9**, **10**, and **11, Additional File 1** for box and locus zoom plots of rs10935473, rs3804622, rs844159, and rs507666, respectively.

Finally, we identified *cis*-pQTLs for CD73: rs2229523 (chr6:86199233; G>A; AAF = 0.34; *P*-value < 2.3e-15), a non-synonymous variant in *NT5E*, the gene that codes CD73, and an intergenic variant, rs494688 (chr6:86100089; G>A; AAF = 0.11; *P*-value < 1.9e-09), 59,712 bases upstream from *NT5E*. The estimated R^2^ between rs2229523 and rs494688 was 0.04 in the analysis data set. See **Table 2**, and **Supplementary Figures 12** and **13, Additional File 1** for box and locus zoom plots of rs2229523, and rs494688, respectively.

### Validation of the association between rs11118119 and TGF-β2 levels

In order to validate the novel *cis*-pQTL for TGF-*β*2 identified in our analysis (rs11118119, chr1:218693872; A>G), we tested the association between rs11118119 and TGF-*β*2 levels in 538 castration-resistant prostate cancer patients from CALGB 90401 and 216 advanced pancreatic cancer patients from CALGB 80303 (13). Selected baseline characteristics for these two cohorts are summarized in **Supplementary Table 3, Additional File 1**.

The association was validated independently in CALGB 90401, where the G allele of rs11118119 (A>G) was associated with higher TGF-*β*2 levels (*P*-value < 3.3e-25, AAF = 0.10), similar to CALGB/SWOG 80405 (**Figure 2**). We could not validate this association in CALGB 80303 (*P*-value = 0.500, AAF = 0.12, **Figure 2**).

### Bioinformatic analysis of rs11118119

Our bioinformatic analysis showed that rs1015275 (G>C), in high LD (R^2^ = 0.91) with rs11118119 (A>G) and located 50 kb downstream from *TGFB2*, is located in the binding motif for HAND1 transcription factor. Additional data from the JASPAR database (40) (using atSNP (37)) predicts preferential binding of HAND1 to the C allele of rs1015275 compared to the G allele (p=0.0038, log-likelihood=-4.46 for the C allele, and p=5.93×10^-6^, log-likelihood=-27.45, p=0.321 for the G allele) (**Supplementary Figure 14, Additional File 1**). This evidence is derived from small-scale in vitro experiments for HAND1 in complex with TCF3 and TCF4. Moreover, the non-coding scores provided by SNPnexus (36) show that rs1015275 has a high predicted functionality according to EIGEN PC (41) (PC score = 1.33) and DeepSEA (42) scores (functional significance score = 0.010). High EIGEN PC scores and low DeepSEA scores indicate that the SNP is predicted to be located in regions of open chromatin, accessible to transcription factors.

## Discussion

The present study investigated the association between genetic markers and circulatory protein levels in mCRC patients and discovered a novel *cis*-pQTL for TGF-*β*2, rs11118119 (A>G) located in *LINC02869*. This finding was supported by further confirmation in an independent external cohort of patients with castration-resistant prostate cancer. Moreover, this study also validated previously discovered *cis*-pQTLs for VEGF-A and CD73, as well as a *trans*-pQTL for VEGF-R3.

The TGF-*β* signaling pathway participates in different biological processes, including cell proliferation, differentiation, adhesion, migration, and apoptosis (43). TGF-*β* acts as a tumor suppressor in normal epithelium cells and in the early stages of different types of cancer, including CRC (44, 45), prostate (46), and pancreatic (47). However, in advanced cancers, TGF-*β* is abundantly expressed and acts as a tumor promoter. The TGF-*β* family consists of three members, TGF-*β*1, TGF-*β*2, and TGF-*β*3. Both TGF-*β*1 and TGF-*β*2 control the activity of stromal cells and tumor cells, affecting cancer progression (48, 49). Higher TGF-*β*2 expression is correlated with the prognosis of different types of cancer, mainly CRC. Higher TGF-*β*2 expression has also been associated with lymph node metastasis in CRC patients and with the expression of several markers of immune cell subspecies in tumors. Thus, TGF-*β*2 expression is related to the magnitude of the tumor infiltration by immune cells, with the potential to serve as a prognostic biomarker in CRC (50).

We provided evidence of replication of the association between rs11118119 and TGF-*β*2 levels in metastatic castration-resistant prostate cancer patients from the CALGB 90401 study. Similar to mCRC patients in CALGB/SWOG 80405, the G allele of rs11118119 (A>G) was associated with higher levels of TGF-*β*2 (**Figure 2**). The association was not replicated for advanced pancreatic cancer patients from CALGB 80303 (**Figure 2**).

The empirical relative allelic frequencies for the risk allele of rs11118119 in the genetically estimated European cohort of CRC, prostate cancer, and pancreatic cancer patients in our study are 0.11, 0.09, and 0.12, respectively. The corresponding reported putative relative frequency in the European (EUR) cohort from the 1000 Genomes database is 0.14 compared to a putative relative frequency of 0.56 in the African (AFR) cohort (51). Effectively, the putative risk allele for this variant is the major allele in the latter population, and this finding might impact a significant proportion of patients with advanced tumors. Bioinformatic analyses showed that rs1015275 (G>C), a SNP in high LD with rs11118119 (A>G) located around 50 kb downstream from *TGFB2*, has high EIGEN PC score and low DeepSEA score, which indicated that the SNP is predicted to be located in regions of open chromatin that are accessible to many transcription factors. Moreover, data from JASPAR database shows that rs1015275 (G>C) is predicted to alter HAND1 binding motif, with the C allele increasing the likelihood of HAND1 binding compared to the G allele. In addition, JASPAR database also shows that HAND1 can complex with TCF3 and TCF4. HAND1, TCF3, and TCF4 are transcription factors of the basic helix-loop-helix protein (bHLH) family, which bind to a consensus sequence, CAnnTG, that resides in *cis*-regulatory elements of downstream target genes (52). Transcription factor interplay is intrinsically related to enhancer function (53), which might indicate higher *TGFB2* expression in patients with the C allele of rs1015275 (corresponding to the G allele of rs11118119), leading to higher circulating levels of TGF-*β*2.

The results of the present investigation validate one of our previous findings that identified rs7767396 as a *cis*-pQTL for circulating VEGF-A in patients with locally advanced pancreatic cancer from CALGB 80303 and in CRC patients in CALGB 80203 (13). From the previous study, it is already known that the binding of NF-AT1 and ZBRK1 transcription factors may be altered by the presence of the G allele of rs7767396 (A>G), which can regulate VEGF-A plasma levels. Moreover, rs7767396, and SNPs in high LD with it (R^2^ >0.95, rs78355601, rs4513773, rs11757903), have been previously associated with VEGF-A plasma levels in several studies reported in the NHGRI-EBI genome-wide association studies (GWAS) catalog (54, 55, 56, 57, 58, 59, 60).

The results of this study also validated previously reported *trans*-pQTLs for VEGF-R3 on chromosomes 3 and 9. On chromosome 3, rs10935473 (C>A) has already been associated with plasma levels of VEGF-R3 in previous studies in patients with pre-diabetes or diabetes reported in the pGWAS database (61) and other studies reported in the NHGRI-EBI GWAS catalog (54). Similar to our study, the A allele of rs10935473 (C>A) was associated with decreased levels of VEGF-R3. On chromosome 9, rs507666 (G>A) has also been associated with plasma levels of VEGF-R3 in a previous study in patients with pre-diabetes or diabetes reported in the pGWAS database (61). Similar to our study, the A allele of rs507666 (G>A) was associated with lower levels of VEGF-R3.

Lastly, our analysis identified rs2229523 (G>A) as a *cis*-pQTL for CD73, with the A allele of rs2229523 in *NT5E* associated with higher plasma levels of CD73. The G allele of rs2229523 (G>A) was already reported as an eQTL decreasing the mRNA expression of *NT5E* in whole blood (p=1.1×10^-5^, normalized effect size NES = −0.16) and many other tissues (62). However, this is the first study reporting rs2229523 as a pQTL for the circulatory protein levels of CD73 in plasma.

This study has some limitations. The discovery process was limited to genetically estimated Europeans. The reported association between rs11118119 and TGF-*β*2 observed in CALGB 80405 (advanced mCRC) and validated in CALGB 90401 (advanced prostate cancer) failed to validate in CALGB 80303 (advanced pancreatic cancer). We note that the TGF-*β*2 assay used in CALGB 80405 and 90401 was an improved version of the assay initially used in CALGB 80303. The first-generation TGF-*β*2 assay did not have as wide a dynamic range or level of sensitivity as the current TGF-*β*2 assay. Further, the TGF-*β*2 assay used in CALGB 80303 had much lower precision, exhibiting a coefficient of variation (CVs) of 15.2% compared to 6.0% and 3.8% observed in CALGB 80405 and CALGB 90401, respectively. The present analysis has been restricted to high quality typed variants at the genotype level. Imputation-and haplotype-based analyses may identify additional relevant sources of genetic variation. Finally, the mechanism proposed of how rs11118119 regulates the levels of TGF-*β*2 by bioinformatic analysis needs to be further validated in experimental models.

## Conclusions

In summary, this study has provided evidence of a novel *cis* germline genetic variant that regulates circulating TGF-*β*2 levels in plasma of patients with advanced CRC and prostate cancer. The putative reference relative allelic frequency for this variant ranges from 0.14 in the European population to over 0.5 in the African population. The discovery of a genetic variant that regulates the levels of TGF-*β*2 in circulation might have important implications for identification of prognostic biomarkers and mechanisms that shape disease heterogeneity in advanced tumors.

## Data Availability

The genotype and phenotype (clinical and protein) data for the CALGB 80405 discovery cohort are are available from the database of Genotypes and Phenotypes (dbGaP) through study accession: phs003428.v1.p1. The data for the CALGB 80303 and CALGB 90401 validation cohorts are available from the database of Genotypes and Phenotypes (dbGaP) through study accession: phs000250.v1.p1 and phs001002.v1.p1 respectively. The TGFB-2 protein markers for the CALGB 80303 are available as supplementary material in Innocenti et al. [1]. The TGFB-2 protein markers for CALGB 90401 will be deposited into dbGaP (phs001002.v1.p1) and are currently available by request from the corresponding author.
[1] Innocenti F, Jiang C, Sibley AB, Etheridge AS, Hatch AJ, Denning S, Niedzwiecki D, Shterev ID, Lin J, Furukawa Y, Kubo M, Kindler HL, Auman JT, Venook AP, Hurwitz HI, McLeod HL, Ratain MJ, Gordan R, Nixon AB, Owzar K. Genetic variation determines VEGF-A plasma levels in cancer patients. Sci Rep. 2018 Nov 5;8(1):16332. doi: 10.1038/s41598-018-34506-4. PMID: 30397360; PMCID: PMC6218528.

## List of abbreviations

AFR: African cohort from the 1000 Genomes database
CALGB: Cancer and Leukemia Group B
VEGF: vascular endothelial growth factor
CONSORT: Consolidated Standards of Reporting Trials
CV: coefficient of variation
EGFR: epidermal growth factor receptor
ELISA: enzyme-linked immunosorbent assay
EUR: European cohort from the 1000 Genomes database
GWAS: genome-wide association study
AAF: alternate allele frequency
mCRC: metastatic colorectal cancer
pQTL: protein quantitative trait loci
QC: quality control
QQ: quantile-quantile
SNP: single nucleotide polymorphism
SWOG: Southwest Oncology Group

## Declarations

## Ethics approval and consent to participate

The analyses were conducted using clinical, protein marker and germline DNA genotyping data from patients registered to CALGB/SWOG 80405 (registered under National Clinical Trial number NCT00265850) who had consented to participation in Institutional Review Board (IRB) approved pharmacogenomic substudy (CALGB 60501). The germline DNA and clinical data used in the validation analyses were obtained from patients from CALGB 90401 (NCT00110214) and CALGB 80303 (NCT00088894) who had provided consent to research under IRB approved pharmacogenomic substudies (CALGB 60404 and CALGB 60401 respectively). CALGB/SWOG 80405, CALGB 90401 and CALGB 80303 were multi-center clinical trials. Each of these three substudies were approved by the Cancer Treatment Evaluation Program (CTEP) of the National Cancer Institute (NCI) and the Institutional Review Board (IRB) of each participating center. The analyses presented in this paper were conducted under protocol Pro00113199 approved by the Duke Health IRB of Duke University.

## Consent for publication

Not applicable.

## Availability of data and materials

The code base to reproduce the statistical and replication analyses presented in this paper is available from a public source code repository (https://gitlab.oit.duke.edu/dcibioinformatics/pubs/calgb-80405-pqtl). The genotype and phenotype (clinical and protein) data for the CALGB 80405 discovery cohort are are available from the database of Genotypes and Phenotypes (dbGaP) through study accession: phs003428.v1.p1. The data for the CALGB 80303 and CALGB 90401 validation cohorts are available from the database of Genotypes and Phenotypes (dbGaP) through study accession: phs000250.v1.p1 and phs001002.v1.p1 respectively. The TGFB-2 protein markers for the CALGB 80303 are available as supplementary material in Innocenti et al. (13). The TGFB-2 protein markers for CALGB 90401 will be deposited into dbGaP (phs001002.v1.p1) and are currently available by request from the corresponding author.

## Authors’ contributions

F.I. and K.O. conceived and conceptualized the study; F.I., J.C.F.Q. and K.O. interpreted the results, and wrote the paper; K.O. led the statistical considerations; A.B.S., L.R. and J.C.F.Q. performed the statistical analyses; J.C.F.Q. performed the bioinformatics analysis; A.B.S and L.R. set up a reproducible analysis workflow and performed the technical tasks for the CALGB 80405 dbGaP data deposit; A.B.N. and Y.L. generated the plasma markers; A.V., B.O. and D.N. designed and wrote the clinical protocol for CALGB 80405; H.K. and D.N. designed and wrote the clinical protocol for CALGB 80303; W.K. and S.H. designed and wrote the clinical protocol for CALGB 90401 clinical trial; A.V. served as study chair for CALGB 80405; H.K. served as study chair for CALGB 80303; W.K. served as study chair for CALGB 90401; D.N. served as lead statistician for CALGB 80303 and 80405; S.H. served as lead statistician for CALGB 90401; H.L.M. and M.J.R. provided resources for the study, and critically reviewed and edited the paper; A.B.S., L.R., S.H. and Y.L. critically edited the paper; All co-authors have read and approved the paper.

## Acknowledgements

The authors thank Dr. Michiaki Kubo of the RIKEN Center for Genomic Medicine of Japan for providing genotyping support for DNA samples from CALGB 80303, 90401 and 80405. The content is solely the responsibility of the authors and does not necessarily represent the official views of the National Institutes of Health.

