## Supplementary Material for "Common variation in a long non-coding RNA gene modulates variation of circulating TGF-*β*2 levels in metastatic colorectal cancer patients (Alliance)"

**Supplementary Table 1. Results and annotation for pQTL that passed the genome-wide threshold.** Prot./Chr: protein marker/chromosome of the variant according to hg19; rsID: RefSNP ID of variant; N: number of samples for whom the variant was called; AAF: relative allelic frequency for the alternate allele; HWE: Hardy-Weinberg *P*-value; Pvalue: *P*-value for pQTL analysis; Est: parameter estimate and 95% CI for the variant effect from the rank-based linear regression; P.c: *P*-value for the variant effect from the rank-based linear regression adjusting for covariates; Est.c: parameter estimate and 95% CI for the variant effect from the rank-based linear regression adjusting for covariates. Variants are sorted by rsID within Protein and Chr. Note that the genotyping data included rs3812138, but this ID has been merged with rs2229523.

| Prot./ Chr | rsID | N | AAF | HWE | Pvalue | Est | P.c | Est.c |
|---|---|---|---|---|---|---|---|---|
| CD73 | | | | | | | | |
| chr6 | rs12212560 | 866 | 0.41 | 0.89 | 8.3E-12 | 0.43 (0.31, 0.55) | 2.2E-12 | 0.43 (0.31, 0.55) |
| | rs1414201 | 869 | 0.33 | 0.54 | 2.3E-09 | 0.41 (0.28, 0.53) | 2.2E-10 | 0.41 (0.29, 0.54) |
| | rs2229523 | 869 | 0.34 | 0.82 | 2.3E-15 | 0.52 (0.40, 0.64) | 9.8E-17 | 0.52 (0.40, 0.65) |
| | rs2842608 | 869 | 0.37 | 0.56 | 2.8E-14 | 0.49 (0.37, 0.61) | 1.1E-15 | 0.50 (0.38, 0.62) |
| | rs4593336 | 869 | 0.37 | 0.38 | 2.7E-12 | 0.44 (0.32, 0.56) | 6.3E-13 | 0.44 (0.32, 0.56) |
| | rs494688 | 869 | 0.11 | 0.73 | 1.9E-09 | -0.61 (-0.80, -0.42) | 8.1E-10 | -0.60 (-0.79, -0.41) |
| | rs6922 | 869 | 0.38 | 0.39 | 1.5E-14 | 0.50 (0.38, 0.62) | 9.5E-16 | 0.50 (0.38, 0.62) |
| | rs6931295 | 865 | 0.34 | 0.94 | 1.3E-13 | 0.49 (0.36, 0.61) | 1.2E-14 | 0.49 (0.37, 0.61) |
| | rs7752502 | 869 | 0.41 | 0.89 | 3.9E-12 | 0.44 (0.32, 0.56) | 1.2E-12 | 0.44 (0.32, 0.55) |
| | rs9362210 | 869 | 0.27 | 0.86 | 1.4E-11 | 0.48 (0.34, 0.61) | 3.4E-12 | 0.47 (0.34, 0.61) |
| | rs9444361 | 869 | 0.38 | 0.52 | 1.1E-13 | 0.48 (0.36, 0.60) | 5.3E-15 | 0.49 (0.37, 0.60) |
| TGF- $\beta$2 | | | | | | | | |
| chr1 | rs11118119 | 868 | 0.11 | 0.48 | 1.4E-14 | 0.52 (0.41, 0.63) | 3.6E-18 | 0.51 (0.40, 0.62) |
| | rs12034138 | 868 | 0.11 | 0.48 | 1.8E-14 | 0.52 (0.40, 0.63) | 3.7E-18 | 0.51 (0.40, 0.62) |
| | rs6665024 | 867 | 0.10 | 0.56 | 3.3E-12 | 0.49 (0.37, 0.61) | 3.4E-15 | 0.48 (0.37, 0.60) |
| | rs725033 | 869 | 0.12 | 0.74 | 1.6E-10 | 0.41 (0.30, 0.52) | 2.1E-12 | 0.40 (0.29, 0.51) |
| VEGF-A | | | | | | | | |
| chr6 | rs7767396 | 868 | 0.48 | 0.95 | 2.6E-12 | -0.51 (-0.66, -0.37) | 9.3E-12 | -0.51 (-0.65, -0.36) |
| | rs9369434 | 868 | 0.43 | 0.78 | 1.2E-09 | -0.45 (-0.60, -0.31) | 3.8E-09 | -0.44 (-0.58, -0.29) |
| | rs9472159 | 868 | 0.49 | 0.59 | 3.8E-11 | -0.48 (-0.62, -0.34) | 2.2E-10 | -0.47 (-0.61, -0.33) |
| VEGF-R3 | | | | | | | | |

| Prot./ Chr | rsID | N | AAF | HWE | Pvalue | Est | P.c | Est.c |
|---|---|---|---|---|---|---|---|---|
| chr3 | rs1000003 | 868 | 0.16 | 0.26 | 6.9E-10 | 0.16 (0.11, 0.22) | 2.2E-08 | 0.16 (0.11, 0.22) |
| | rs10935473 | 869 | 0.45 | 0.49 | 6.4E-39 | -0.29 (-0.33, -0.25) | 1.4E-43 | -0.29 (-0.33, -0.25) |
| | rs1531377 | 869 | 0.17 | 0.72 | 4.1E-10 | 0.16 (0.11, 0.22) | 1.2E-08 | 0.16 (0.11, 0.22) |
| | rs1675526 | 865 | 0.32 | 0.94 | 3.6E-15 | 0.18 (0.13, 0.22) | 6.7E-15 | 0.18 (0.14, 0.23) |
| | rs3755569 | 869 | 0.27 | 0.10 | 4.5E-11 | 0.15 (0.10, 0.20) | 3.5E-10 | 0.15 (0.10, 0.20) |
| | rs3804622 | 869 | 0.51 | 0.46 | 5.7E-23 | -0.21 (-0.25, -0.17) | 2.0E-24 | -0.22 (-0.26, -0.18) |
| | rs4558798 | 869 | 0.32 | 0.76 | 2.6E-15 | 0.18 (0.13, 0.23) | 4.2E-15 | 0.18 (0.14, 0.23) |
| | rs4857412 | 869 | 0.15 | 0.18 | 1.1E-09 | 0.17 (0.11, 0.23) | 1.3E-08 | 0.17 (0.11, 0.23) |
| | rs4857414 | 869 | 0.47 | 0.45 | 1.7E-32 | -0.26 (-0.30, -0.22) | 2.2E-35 | -0.26 (-0.30, -0.22) |
| | rs6797163 | 869 | 0.34 | 0.82 | 3.3E-20 | 0.20 (0.16, 0.25) | 4.6E-19 | 0.20 (0.16, 0.25) |
| | rs7103 | 869 | 0.36 | 0.27 | 1.3E-10 | -0.14 (-0.19, -0.10) | 4.2E-11 | -0.15 (-0.19, -0.10) |
| | rs7628381 | 869 | 0.27 | 0.10 | 3.1E-11 | 0.15 (0.10, 0.20) | 2.5E-10 | 0.15 (0.11, 0.20) |
| | rs7630058 | 869 | 0.23 | 0.45 | 2.3E-11 | 0.16 (0.11, 0.21) | 7.1E-10 | 0.16 (0.11, 0.21) |
| | rs7653204 | 866 | 0.16 | 0.26 | 9.2E-10 | 0.16 (0.11, 0.22) | 2.5E-08 | 0.16 (0.11, 0.22) |
| | rs844159 | 869 | 0.46 | 0.58 | 1.1E-19 | 0.20 (0.15, 0.24) | 5.9E-20 | 0.20 (0.16, 0.24) |
| | rs865474 | 868 | 0.46 | 0.63 | 3.3E-19 | 0.19 (0.15, 0.23) | 2.2E-19 | 0.19 (0.15, 0.23) |
| | rs9825798 | 869 | 0.17 | 0.64 | 2.0E-10 | 0.17 (0.11, 0.22) | 6.0E-09 | 0.17 (0.11, 0.22) |
| chr9 | rs507666 | 869 | 0.20 | 0.17 | 9.1E-12 | -0.19 (-0.24, -0.13) | 2.3E-12 | -0.19 (-0.25, -0.14) |
| | rs579459 | 869 | 0.23 | 0.08 | 9.2E-12 | -0.18 (-0.23, -0.12) | 1.3E-11 | -0.18 (-0.23, -0.13) |

**Supplementary Table 2. Additional annotation for pQTL that passed the genome-wide threshold**. Protein: protein marker; rsID: refSNP ID of variant; Chr: chromosome of the variant according to hg19; Pos: of the variant according to hg19; Base: Illumina TOP reference>alternate alleles; Gene: gene symbol for intragenic variants or nearest downstream/upstream gene symbols for intergenic variants; Anno: functional annotation according to SNP Nexus. Note that *C1orf143* is an alias for *LINC02869*.

| Protein/rsID | Chr | Pos | Base | Gene | Anno |
|---|---|---|---|---|---|
| CD73 | | | | | |
| rs12212560 | chr6 | 86342679 | G>A | SYNCRIP | intronic |
| rs1414201 | chr6 | 86500316 | G>A | RP11-207F8.1/ Y_RNA | |
| rs2229523 | chr6 | 86199233 | G>A | NT5E | coding nonsyn, coding *nonsyn |
| rs2842608 | chr6 | 86295858 | C>A | SNX14, RP11-321N4.5 | intronic |
| rs4593336 | chr6 | 86181931 | G>A | NT5E | 3downstream, intronic |
| rs494688 | chr6 | 86100089 | G>A | DUTP5/ RP11-30P6.6 | |
| rs6922 | chr6 | 86205323 | C>A | NT5E | 3utr |
| rs6931295 | chr6 | 86144875 | G>A | NT5E/ TPT1P6 | |
| rs7752502 | chr6 | 86410651 | A>G | RP11-33E24.3/ SNHG5 | |
| rs9362210 | chr6 | 86118728 | G>A | DUTP5/ RP11-30P6.6 | |
| rs9444361 | chr6 | 86400900 | A>G | RP11-33E24.3/ SNHG5 | |
| TGF-b2 | | | | | |
| rs11118119 | chr1 | 218693872 | A>G | C1orf143 | intronic |
| rs12034138 | chr1 | 218692641 | G>A | C1orf143 | intronic |
| rs6665024 | chr1 | 218690577 | G>A | C1orf143 | intronic |
| rs725033 | chr1 | 218643940 | G>A | C1orf143/ TGFB2 | |
| VEGF-A | | | | | |
| rs7767396 | chr6 | 43927050 | A>G | RP5-1120P11.1/ RP11-344J7.2 | |
| rs9369434 | chr6 | 43918407 | G>A | RP5-1120P11.1/ RP11-344J7.2 | |
| rs9472159 | chr6 | 43919695 | C>A | RP5-1120P11.1/ RP11-344J7.2 | |
| VEGF-R3 | | | | | |
| rs1000003 | chr3 | 98342907 | A>G | WWP1P1/ AC021660.1 | |
| rs10935473 | chr3 | 98416900 | C>A | ST3GAL6-AS1/ WWP1P1 | |
| rs1531377 | chr3 | 98343242 | G>A | WWP1P1/ AC021660.1 | |
| rs1675526 | chr3 | 98319101 | A>G | RP11-227H4.1/ CPOX | |
| rs3755569 | chr3 | 98449742 | G>A | ST3GAL6-AS1 | non-coding intronic |
| rs3804622 | chr3 | 98303182 | G>A | CPOX | intronic, non-coding intronic |
| rs4558798 | chr3 | 98308915 | G>A | CPOX | intronic, 3downstream |
| rs4857412 | chr3 | 98391040 | A>G | ST3GAL6-AS1/ WWP1P1 | |
| rs4857414 | chr3 | 98432559 | G>A | ST3GAL6-AS1/ WWP1P1 | |
| rs6797163 | chr3 | 98367525 | G>A | WWP1P1/ AC021660.1 | |
| rs7103 | chr3 | 98298600 | G>A | CPOX | 3utr, intronic, 3downstream |
| rs7628381 | chr3 | 98442701 | G>A | ST3GAL6-AS1 | non-coding intronic |
| rs7630058 | chr3 | 98337517 | G>A | RP11-227H4.1/ CPOX | |
| rs7653204 | chr3 | 98346506 | A>G | WWP1P1/ AC021660.1 | |
| rs844159 | chr3 | 98443648 | A>G | ST3GAL6-AS1 | non-coding intronic |
| rs865474 | chr3 | 98445324 | A>G | ST3GAL6-AS1 | non-coding intronic |
| rs9825798 | chr3 | 98385816 | A>G | ST3GAL6-AS1/ WWP1P1 | |
| rs507666 | chr9 | 136149399 | G>A | ABO | non-coding intronic |
| rs579459 | chr9 | 136154168 | A>G | Y_RNA/ ABO | |

**Supplementary Table 3. Demographics and clinical characteristics patients of genetically determined European ancestry included in the replication analyses of CALGB 90401 and CALGB 80303.** Prior radiation was not available for CALGB 90401. In CALGB 80303, ECOG Performance Status was only reported as 0 or 1 vs. 2.

| | CALGB 90401<br>n=538 | CALGB 80303<br>n=216 |
|---|---|---|
| **Age decade – n (%)** | | |
| 3 | | 3 (1.39%) |
| 4 | 11 (2.04%) | 14 (6.48%) |
| 5 | 75 (13.9%) | 47 (21.8%) |
| 6 | 211 (39.2%) | 62 (28.7%) |
| 7 | 202 (37.6%) | 55 (25.5%) |
| 8 | 36 (6.7%) | 35 (16.2%) |
| 9 | 3 (0.56%) | |
| **Gender – n (%)** | | |
| Male | 538 (100.0%) | 118 (54.6%) |
| Female | | 98 (45.4%) |
| **Treatment arm – n (%)** | | |
| Bevacizumab | 270 (50.2%) | 111 (51.4%) |
| Placebo | 268 (49.8%) | 105 (48.6) |
| **Prior radiation – n (%)** | | |
| No | | 195 (90.3%) |
| Yes | | 21 (9.7%) |
| **ECOG PS – n (%)** | | |
| 0 | 298 (55.4%) | 195 (90.3%) |
| 1 | 226 (42 %) | 21 (9.7%) |
| 2 | 14 (2.6%) | |

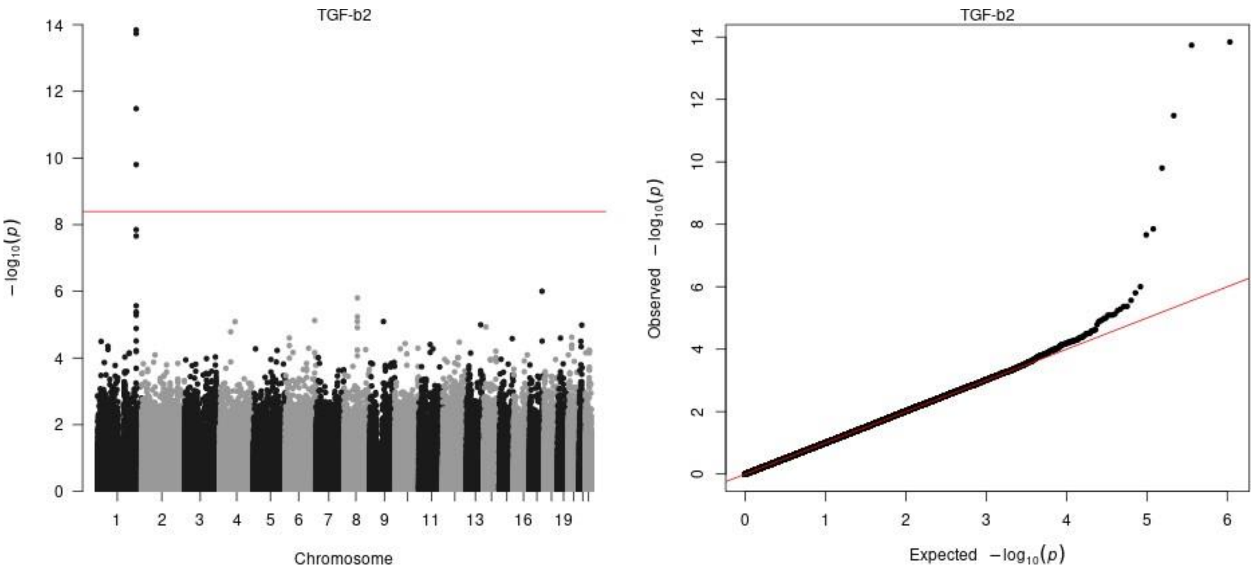


**Supplementary Figure 1.** Manhattan and QQ plots of the empirical unadjusted *P*-values for the pQTL analysis of TGF-$\beta$2.

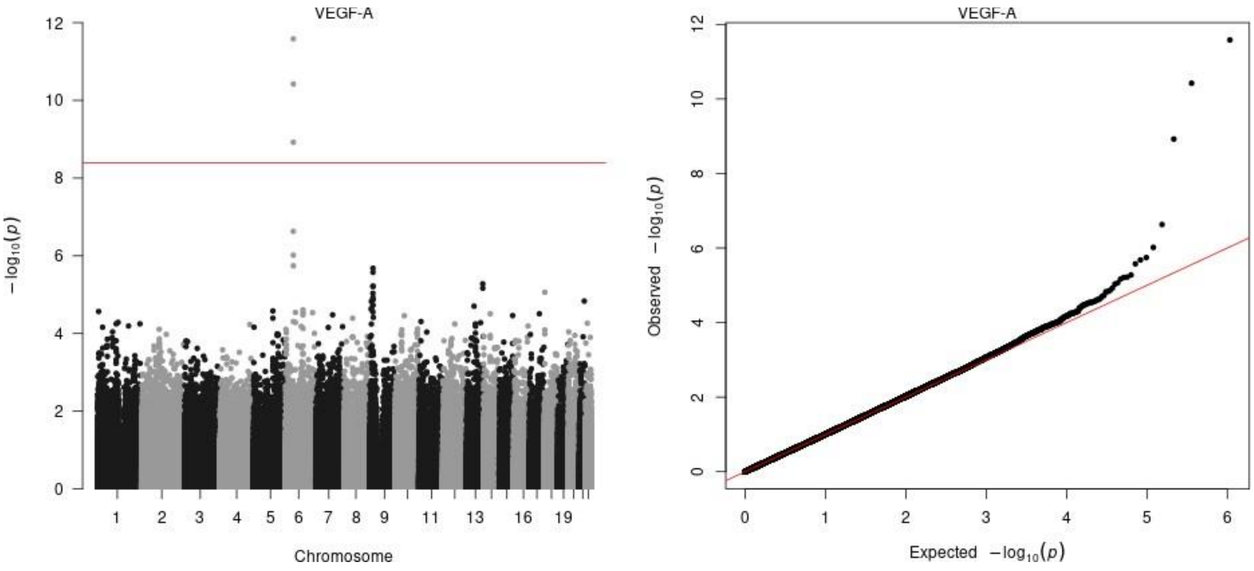


**Supplementary Figure 2.** Manhattan and QQ plots of the empirical unadjusted *P*-values for the pQTL analysis of VEGF-A.

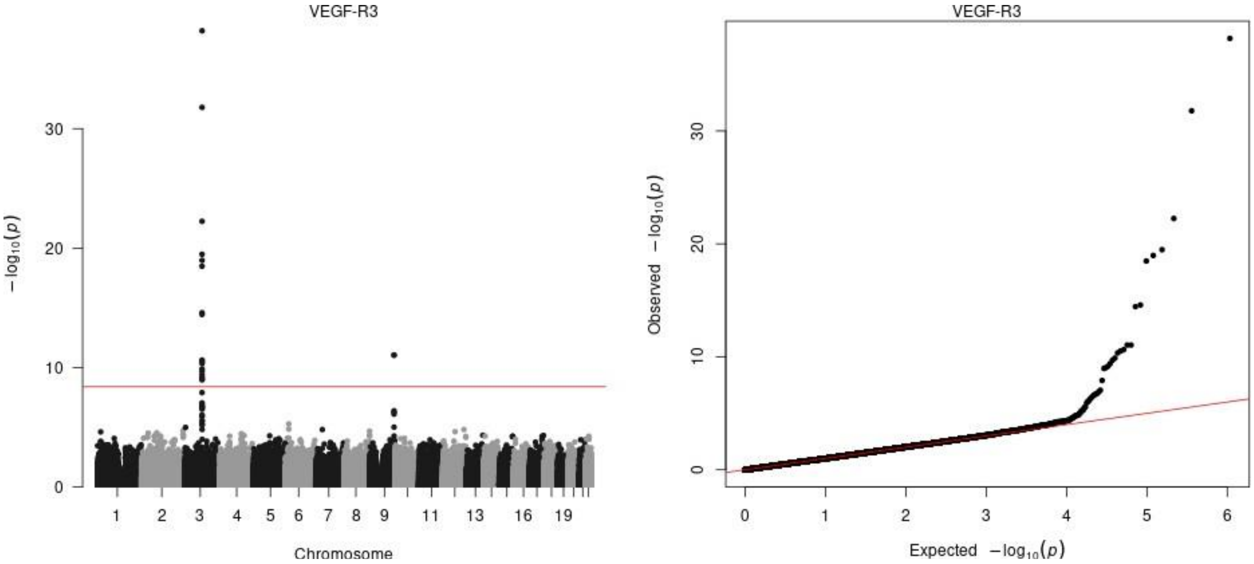


**Supplementary Figure 3.** Manhattan and QQ plots of the empirical unadjusted *P*-values for the pQTL analysis of VEGF-R3

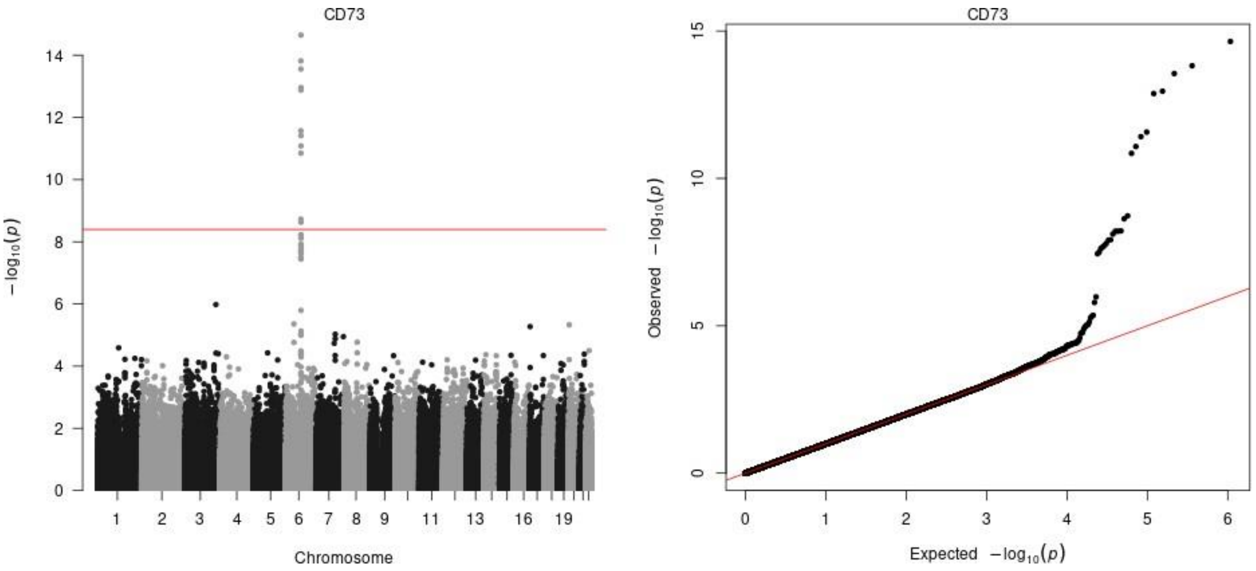


**Supplementary Figure 4.** Manhattan and QQ plots of the empirical unadjusted *P*-values for the pQTL analysis of CD73.

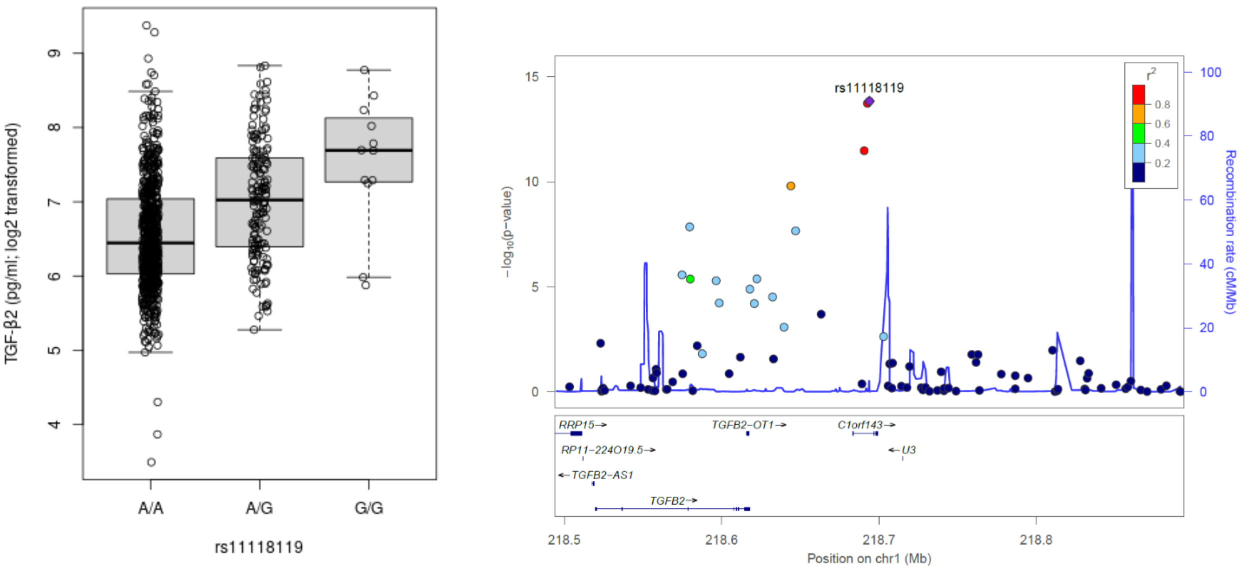


**Supplementary Figure 5.** Boxplot of distributions of log base 2 transformed protein abundances conditional on genotype (left panel) and locus zoom plot (right panel) for pQTL (rs11118119, TGF-$\beta$2).

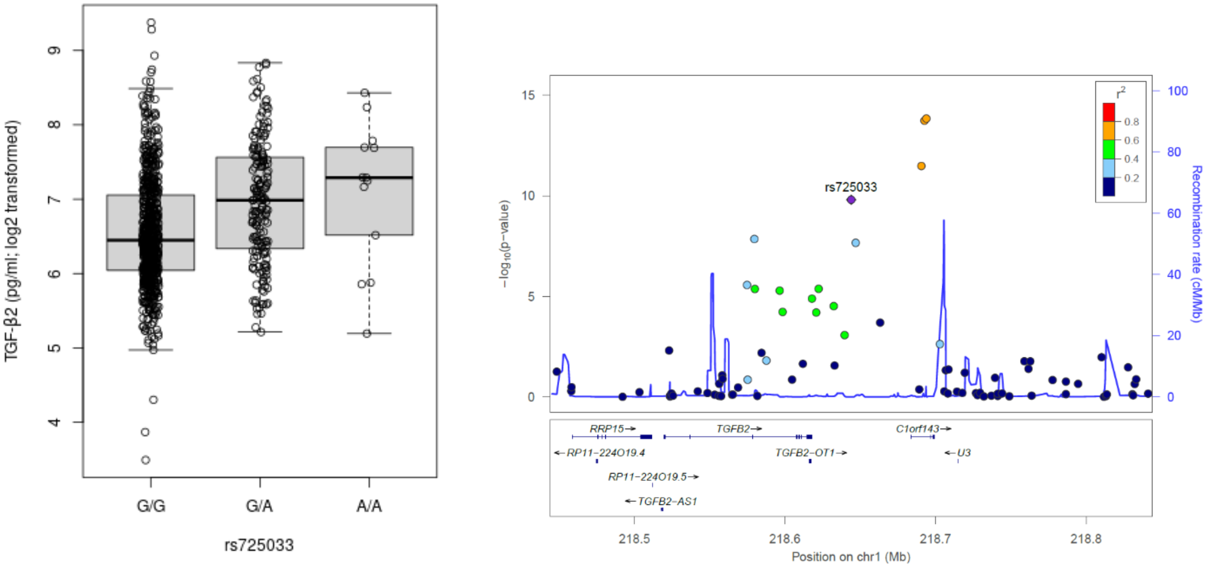


**Supplementary Figure 6.** Boxplot of distributions of log base 2 transformed protein abundances conditional on genotype (left panel) and locus zoom plot (right panel) for pQTL (rs725033, TGF-β2).

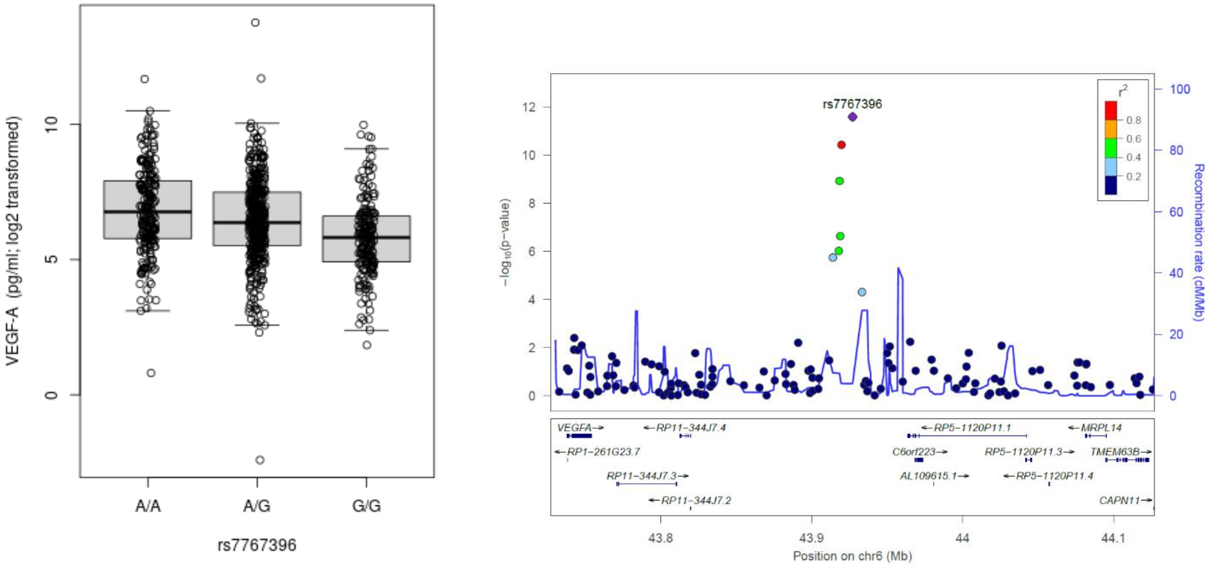


**Supplementary Figure 7.** Boxplot of distributions of log base 2 transformed protein abundances conditional on genotype (left panel) and locus zoom plot (right panel) for pQTL (rs7767396, VEGF-A).

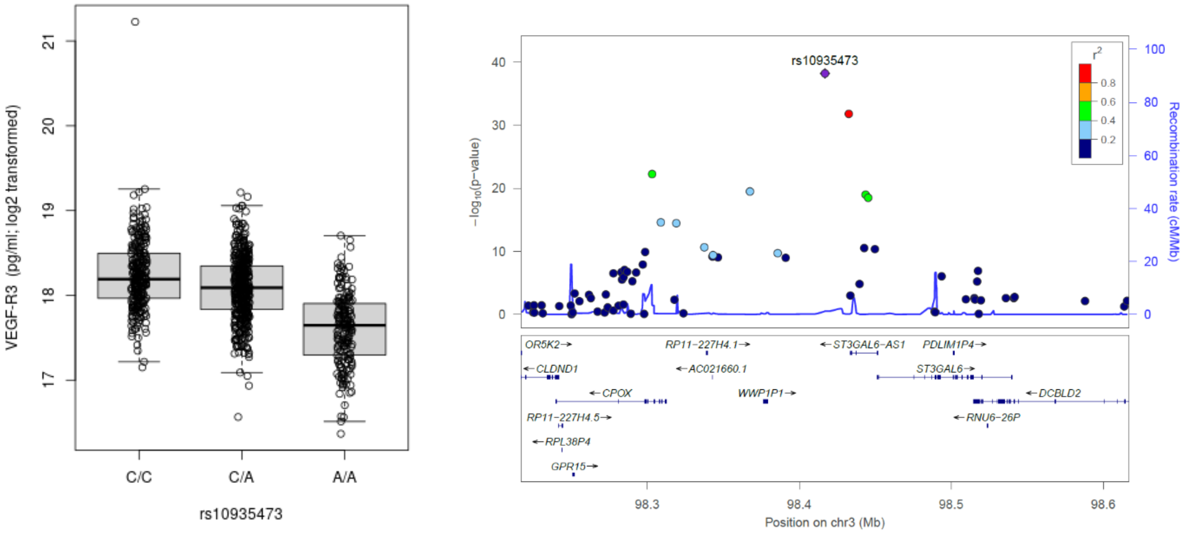


**Supplementary Figure 8.** Boxplot of distributions of log base 2 transformed protein abundances conditional on genotype (left panel) and locus zoom plot (right panel) for pQTL (rs10935473, VEGF-R3).

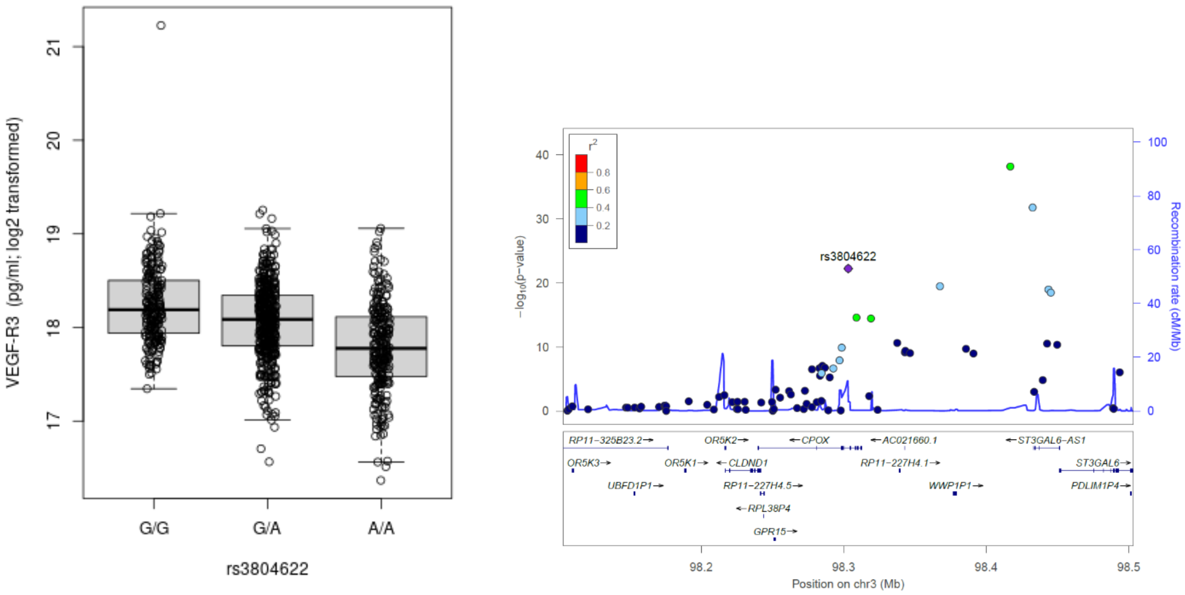


**Supplementary Figure 9.** Boxplot of distributions of log base 2 transformed protein abundances conditional on genotype (left panel) and locus zoom plot (right panel) for pQTL (rs3804622, VEGF-R3).

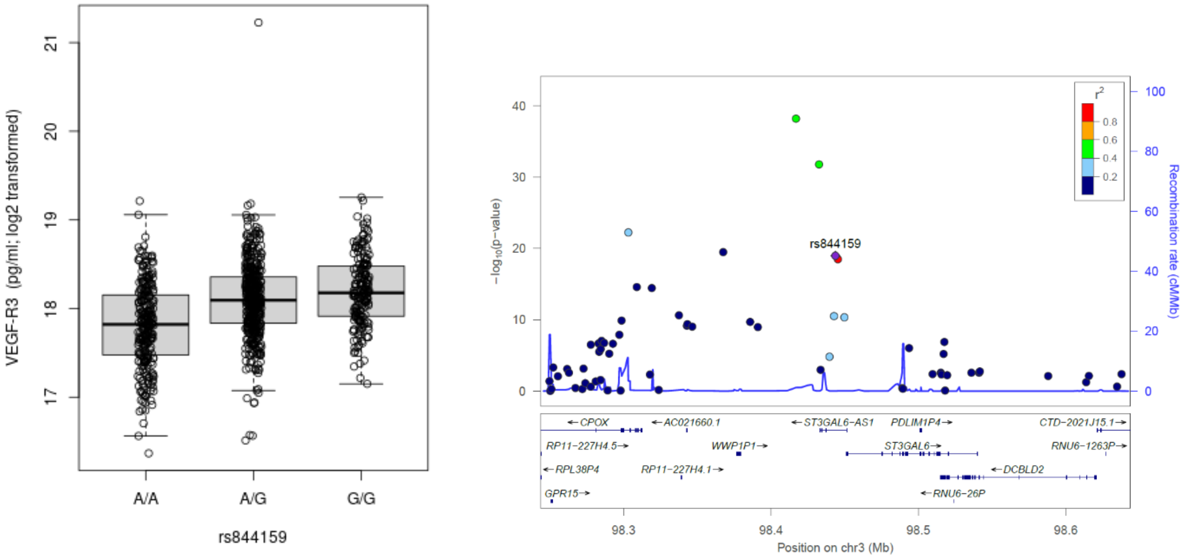


**Supplementary Figure 10.** Boxplot of distributions of log base 2 transformed protein abundances conditional on genotype (left panel) and locus zoom plot (right panel) for pQTL (rs844159, VEGF-R3).

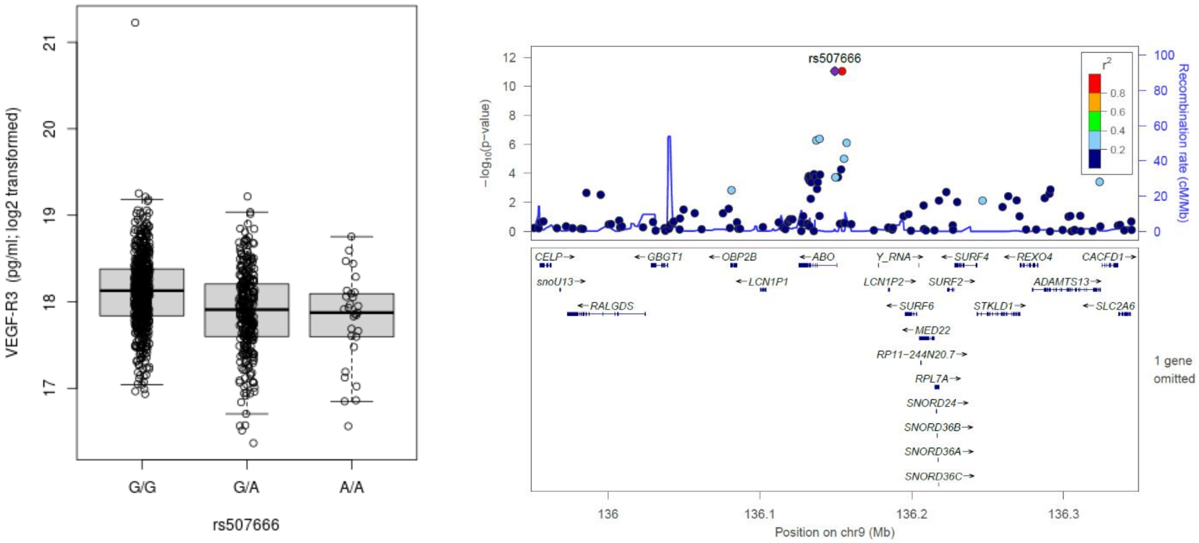


**Supplementary Figure 11.** Boxplot of distributions of log base 2 transformed protein abundances conditional on genotype (left panel) and locus zoom plot (right panel) for pQTL (rs507666, VEGF-R3).

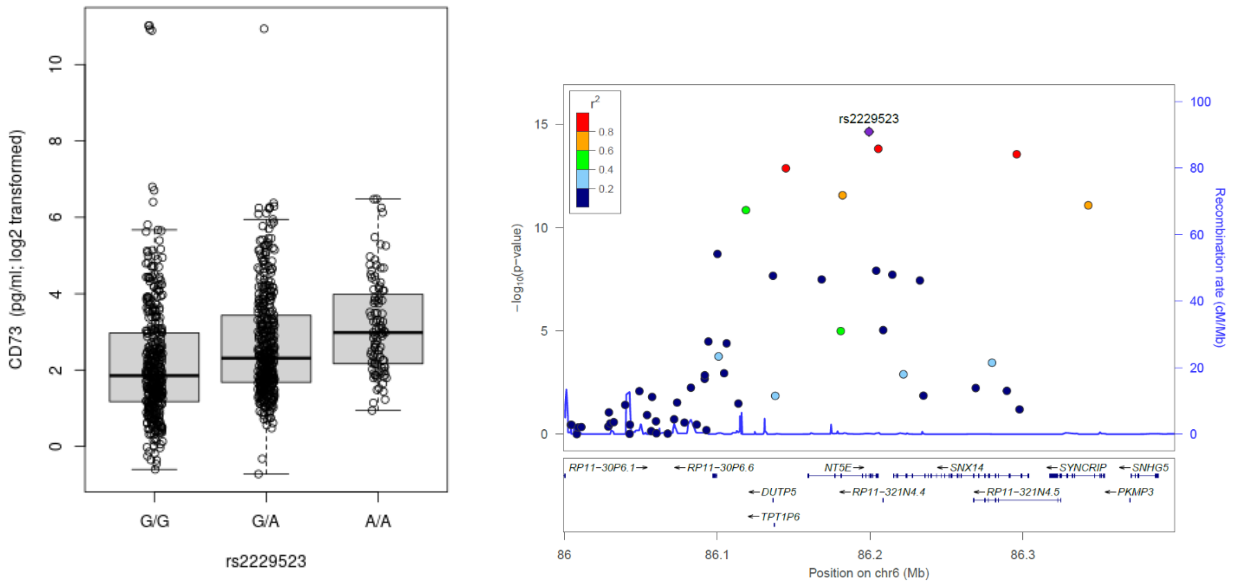


**Supplementary Figure 12.** Boxplot of distributions of log base 2 transformed protein abundances conditional on genotype (left panel) and locus zoom plot (right panel) for pQTL (rs2229523, CD73).

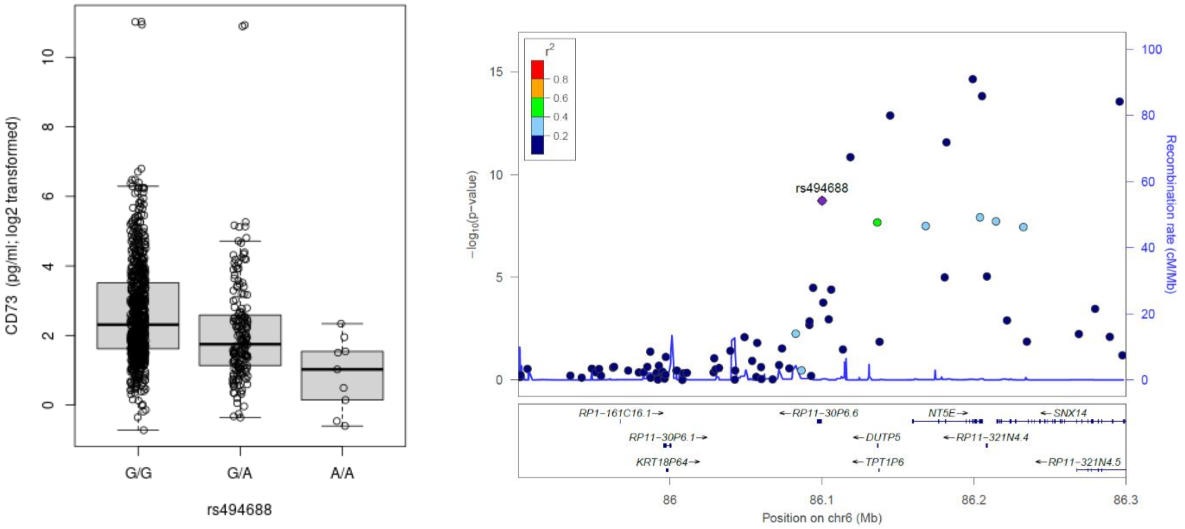


**Supplementary Figure 13**. Boxplot of distributions of log base 2 transformed protein abundances conditional on genotype (left panel) and locus zoom plot (right panel) for pQTL (rs494688, CD73).

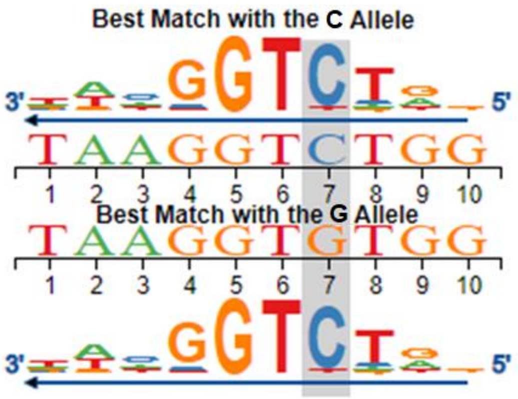


**Supplementary Figure 14. Position of rs1015275 (G>C), 50 kb downstream from *TGFB2*, relative to the HAND1 transcription factor binding motif and effect of the allele change (G versus C).** Sequence logo from atSNP representing the JASPAR positional weight matrix of the HAND1 binding motif. The sequence of the putative regulatory region surrounding rs1015275 (highlighted in gray), with the minor allele (C, top) or reference allele (G, bottom), is shown overlapping the JASPAR HAND1 binding motif. The height of the bases indicates the level of conservation across species, suggesting a regulatory role. rs1015275 results in a nucleotide change from G to C in a conserved region of the HAND1 binding motif.
